# Whole-genome variant detection in long-read sequencing data from ultra-low input patient samples

**DOI:** 10.1101/2025.07.25.25332067

**Authors:** Katherine Wang, Cera J. Aex, Hayan Lee, Lucas Finot, Kevin Zhu, Julianna R. Chang, Aaron M. Horning, William J. Rowell, Philip Li, Sarah B. Kingan, Michael P. Snyder, Graham S. Erwin

**Affiliations:** Department of Molecular and Human Genetics, Baylor College of Medicine, Houston, Texas 77030, USA; Department of Genetics, Stanford University, Palo Alto, California 94304, USA; Cancer Epigenetics Institute, Nuclear Dynamics and Cancer Program, Fox Chase Cancer Center, Philadelphia, Pennsylvania 19111, USA; Pacific Biosciences, Menlo Park, California 94025, USA

## Abstract

Long-read sequencing provides a more complete view of the genome than short-read sequencing, with improved detection of structural variants, tandem repeats, and small variants (single nucleotide variants and insertions and deletions) in difficult-to-map regions. One limitation of long-read sequencing has been high input DNA requirements, with several micrograms required per sample. Here, we evaluate two methods of amplification-based long-read, whole-genome sequencing: ultra-low input HiFi (ULI-HiFi) sequencing and droplet multiple displacement amplification (dMDA) sequencing. When benchmarked against the Genome in a Bottle reference set (NA24385), we observe high precision and recall of single nucleotide variants (SNVs) with ULI-HiFi compared to the dMDA-amplified samples (F1 scores for SNVs of 99.82% for ULI-HiFi compared to 89.46% for dMDA). Across a catalog of >1.6 million tandem repeats (TRs), ULI-HiFi achieves 90.4% perfect concordance and 98.9% accuracy when allowing for single motif differences. ULI-HiFi also illuminates medically-important genes that were poorly mapped by short-read sequencing. We further apply ULI-HiFi to analyze a normal, polyp, and adenocarcinoma sample from a patient with familial adenomatous polyposis (FAP), a hereditary form of colorectal cancer. We identify a TR that progressively expanded in length from normal to polyp to adenocarcinoma. This repeat is located in the 5′ UTR of *LIMD1,* a reported tumor suppressor. Reporter assays reveal significantly reduced expression in colorectal cancer cell lines with increasing repeat length in the *LIMD1* 5’ UTR. We conclude that ULI-HiFi improves the characterization of genetic variants in dark regions of genomes from patient samples, enabling a better understanding of human disease.

## Introduction

With the widespread adoption of whole-genome sequencing, comprehensive genomic analysis has emerged as an increasingly important tool for elucidating the molecular basis of human disease and accelerating precision medicine approaches. Most genome sequencing projects rely on short-read sequencing (SRS), which produces highly accurate reads that are limited in read length (usually 150 bp). With the recent shift from targeted gene panels to SRS-based whole-genome sequencing (WGS), there are hundreds of thousands of regions spanning hundreds of megabases of the genome that are effectively invisible to short reads (Ebbert et al. 2019). In fact, approximately 5–6% of the human genome continue to lack proper alignment and annotation with short read approaches (Vestergaard et al. 2021; Chintalaphani et al. 2021; Mahmoud et al. 2024). Improvements in sequencing methods and new bioinformatic tools have relieved many shortcomings of SRS for variant detection (Dolzhenko et al. 2017; Dashnow et al. 2018; Mousavi et al. 2019; Chen et al. 2016), these methods still struggle to resolve many regions of the genome, namely highly repetitive regions, including segmental duplications and tandem repeats (TRs) larger than the sequencing read length (Olson et al. 2023; Tanudisastro et al. 2024). As a result, many disease-causing mutations, particularly in repetitive regions, are missed with SRS, highlighting a need for more complete, highly accurate genome sequencing technologies (Mandelker et al. 2016; English et al. 2024).

New long-read sequencing technologies, which generate sequencing reads that are thousands of base pairs in length or more, can detect structural variants, phase haplotypes, and genotype SNVs and indels in repetitive regions and regions with high GC content (Chintalaphani et al. 2021; Hu et al. 2021). Long-read sequencing can thus improve genome assembly and small variant detection across the whole genome, including in difficult-to-map regions of the reference genome. PacBio continuous long-read (CLR) sequencing and Oxford Nanopore Technologies (ONT) sequencing both reliably produce long reads but have historically been constrained by reduced read accuracy. PacBio CLR achieves ∼87% accuracy of base calls (SNVs and indels), but ONT-based sequencing has steadily improved, reaching >95% accuracy, with some recent reports suggesting even higher accuracy (Eid et al. 2009; Mikheyev and Tin 2014; Mahmoud et al. 2019; Shafin et al. 2021; Kolesnikov et al. 2024). This high error rate means long-read technologies have rarely been used to detect SNVs or indels. By contrast, PacBio HiFi technology produces highly accurate (>99%) long (10–20 kb) consensus reads from noisy individual subreads (Travers et al. 2010; Loomis et al. 2013; Wenger et al. 2019). In one recent report, these technologies have enabled the genotyping of >200,000 single nucleotide variants (SNVs) and >150,000 insertions and deletions (indels) that were absent from short read analyses (Kronenberg et al. 2025). Despite their benefits, the higher cost of sequencing (approximately 5× the cost of short-read sequencing) (Espinosa et al. 2024), and the need for dedicated equipment—particularly for germline testing in rare diseases—have prevented their widespread adoption in clinical practice. In addition, long-read sequencing methods are also limited by their large (microgram quantity) sample inputs, which frustrates many applications in fields like rare disease screening in newborns and biobanked samples that are limited in quantity.

Impressive advances in high-fidelity DNA amplification have made long-read single-cell sequencing possible for the first time (Fan et al. 2021; Hård et al. 2023). In one study, single CD8+ T cells (∼6 picograms of DNA per cell) were sequenced, enabling detection of single nucleotide variants (SNVs), structural variants (SVs), and tandem repeats (TRs) (Hård et al. 2023). Nevertheless, allelic dropout and amplification errors remain a challenge in these single-cell approaches (Fan et al. 2021; Hård et al. 2023). We believe that an approach that focuses on the amplification of DNA from small (nanogram) amounts of material could yield accurate SNV detection and balanced coverage, which would be clinically useful.

Here, we tested two methods that leverage PacBio’s HiFi sequencing technology to produce high-accuracy long-read data from ultra-low input (ULI) amounts of DNA at the nanogram scale. To identify which amplification method to select, we benchmarked the detection of SNVs, insertions and deletions (indels), structural variants (SVs), and tandem repeats (TRs) against the Genome in a Bottle (GIAB) reference set for NA24385. We then applied ULI-HiFi to investigate small variants, indels, SVs, TRs, and other genetic alterations in clinical samples from a patient with familial adenomatous polyposis (FAP), aiming to uncover somatic structural variants and TR expansions associated with disease development and progression. Overall, this study highlights the potential for long-read HiFi sequencing technology to expand our understanding of human genomics in applications where insufficient DNA yields have traditionally restricted sequencing approaches.

## Results

### Comparison of two methods for HiFi sequencing from ultra low-input samples (ULI)

Standard PacBio HiFi sequencing currently requires several micrograms of DNA input, which poses a significant challenge for many clinical samples. We set out to evaluate amplification-based approaches to sequence whole human genomes. We compared two methods to amplify genomic DNA from ultra-low input (nanogram-level) human genomic DNA (**Fig. 1**).

**Figure 1.**
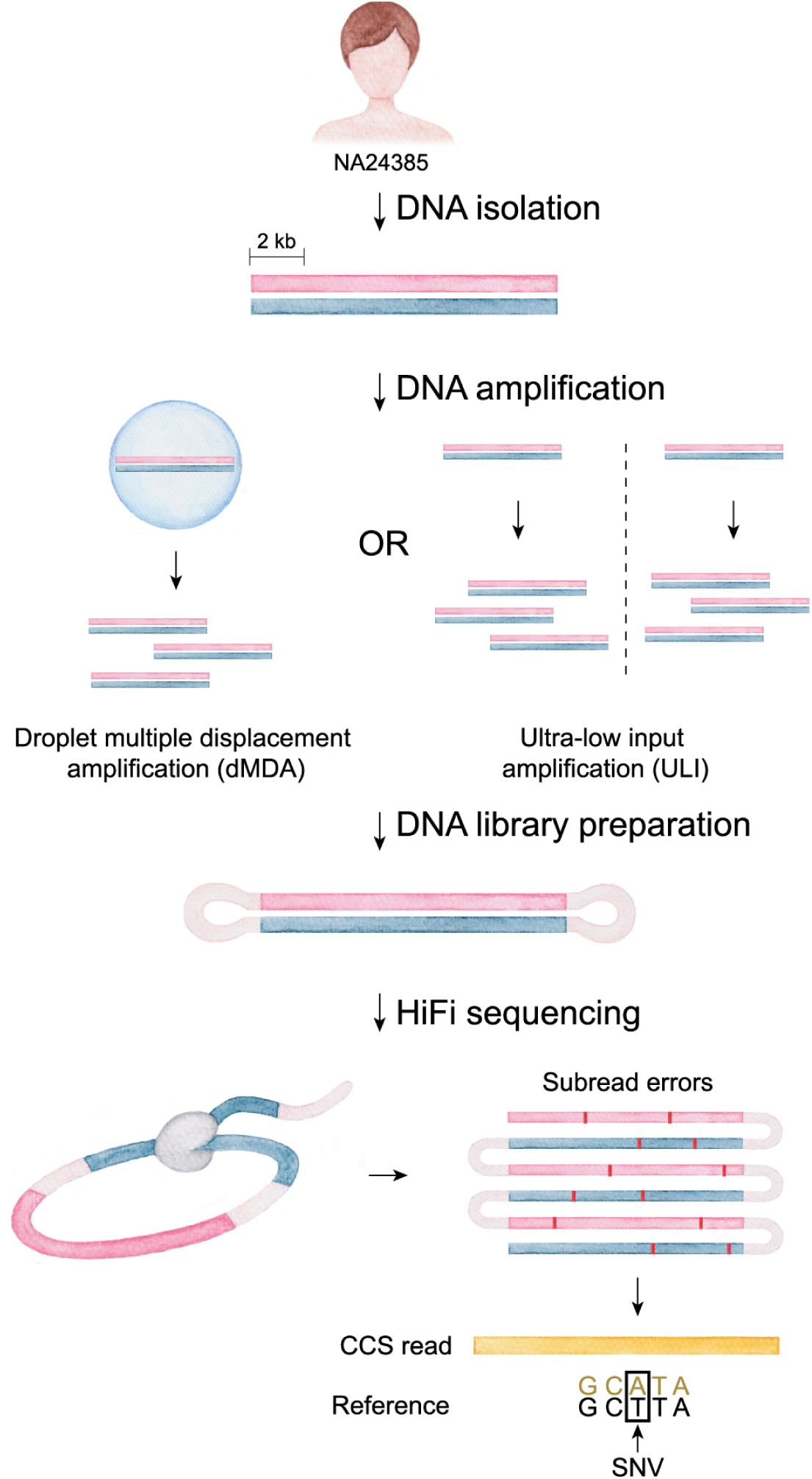
Sequencing NA24385 (HG002) with HiFi long reads from ultra-low input DNA samples. Schematic of the workflow, with two amplification strategies tested. Twenty (20) ng of DNA was amplified using the ultra-low input amplification (ULI) method and 0.45 ng of DNA was amplified using droplet multiple displacement amplification (dMDA), according to manufacturer protocols.

The first strategy, droplet multiple displacement amplification (dMDA), amplifies long (several kb length) DNA molecules in droplets. With dMDA, millions of double-emulsion droplets are created to partition DNA molecules into each droplet with a target of one molecule per droplet. The second strategy is a bulk amplification method, termed ultra-low input (ULI) amplification. The ULI method is a PCR-based method that uses two parallel amplifications to optimize enrichment of both AT- and GC-rich regions to produce a more uniform amplification profile. This protocol is designed to produce more uniform coverage across the genome, particularly in regions with dense GC or AT sequences. This parallel amplification approach minimizes bias previously noted with other amplification-based techniques, particularly with short-read sequencing platforms (Aird et al. 2011; Ross et al. 2013).

We obtained DNA from reference sample NA24385 (HG002), which was selected because it is a broadly consented, extensively studied sample with a reference call set for both small variants (SNVs and indels) and larger SVs from the Genome in a Bottle Consortium (Zook et al. 2020; Wagner et al. 2022). NA24385 DNA was amplified using the dMDA and ULI approaches, followed by standard library preparation for PacBio HiFi sequencing to generate long-read data (see **Methods**). Over the course of our sequencing efforts, the PacBio Revio platform became widely available; therefore, some of our samples were sequenced on the Sequel II platform, while others were done on the Revio platform (**Table 1**). This affected sequencing depth per SMRT cell, but overall quality of sequencing reads at a given depth remains consistent. Additionally, to ensure that observed differences between amplification strategies were not driven by platform-specific effects, we performed a platform benchmarking analysis using NA24385 data from Genome in a Bottle processed through the same alignment and variant-calling workflow (Zook et al. 2016; Wenger et al. 2019). Consistent performance across platforms was observed at matched depth, echoing two previous reports published in this journal (Tesi et al. 2024; Harvey et al. 2023). Our results, combined with those of two other groups, allowed us to conclude that differences in variant calling accuracy are overwhelmingly from the amplification method rather than the sequencing platform (**Supplemental Table S1**). Sequencing was performed using 1 SMRT cell for ULI-HiFi, 4 SMRT cells for dMDA, and 2 SMRT cells for Standard HiFi, resulting in coverage depths of 27.5×, 15.5×, and 28×, respectively (**Table 1**; **Supplemental Table S2**). First, we compared small variants (SNVs, indels <50 bp) data from each approach. Accuracy was determined by comparing the results to the Genome in a Bottle (GIAB) benchmark set of small variants (v4.2.1; **Table 1**).

**Table 1.**
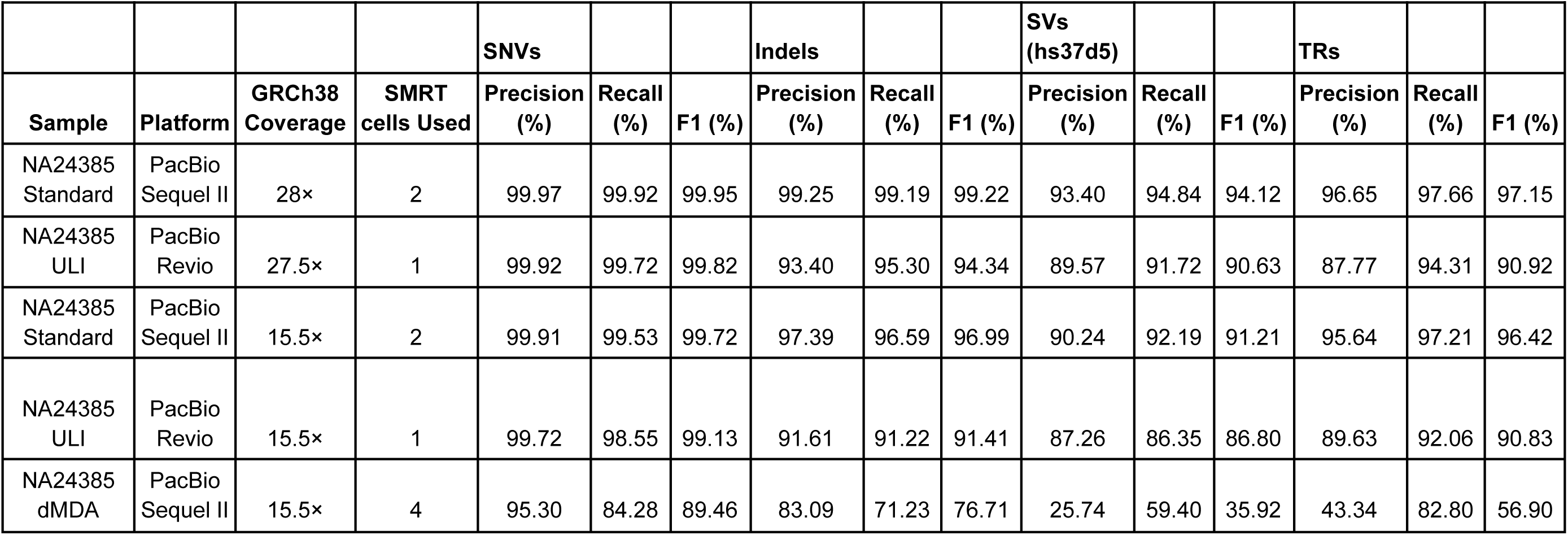
Performance of variant calling in NA24385 (HG002). Performance of small variant calling (SNVs and indels) was from GRCh38 and measured against Genome in a Bottle (GIAB) small variant reference set v4.2.1. Performance of SV calling was from hs37d5 and measured against GIAB SV reference set v0.6. Performance of TR calling was from GRCh38 and benchmarked with GIAB tandem repeat (TR) reference set v1.0.1. As a metric of accuracy, F1 score was calculated as 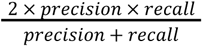, with an F1 score of 100% indicating perfect precision and recall. NA24385 ULI and Standard downsampled to 15.5× are reported as the average of three independently subsampled replicates (see **Methods**). All samples were analyzed with DeepVariant v1.4 (SNVs and indels), Sniffles v2.2 (SVs), and TRGT v1.1.1 (TRs).

In our hands, we found that the accuracy of SNV detection from ULI-HiFi was nearly equivalent to that of Standard HiFi in accuracy of SNV detection (F1 values of 99.82% and 99.95% for ULI-HiFi and Standard HiFi, respectively). The detection of indels performed worse (F1 value of 94.34% and 99.22% for ULI-HiFi and Standard HiFi, respectively). However, when homopolymers were removed from the analysis, indel accuracy improved (F1 value of 96.02%) (**Supplemental Table S3**). By contrast, the accuracy of SNV detection from dMDA HiFi data was substantially lower (F1 values of 89.46% and 76.71% for SNVs and indels, respectively). Even after downsampling the ULI-HiFi data to match the coverage in dMDA, ULI-HiFi still demonstrated high accuracy (F1 values of 99.13% and 91.41% for SNVs and indels, respectively) (**Table 1**). For depth-matched comparisons, we also downsampled the Standard HiFi NA24385 dataset (28×) to 15.5× to match the lowest-coverage dataset (dMDA), and repeated benchmarking against GIAB using the same pipeline. To confirm that depth matching was not sensitive to the specific random seed used for subsampling, we generated three independent 15.5× subsamples for both Standard HiFi and ULI-HiFi NA24385 datasets and benchmarked each replicate against the GIAB benchmark. Benchmark metrics were highly consistent across replicates (coefficient of variation <1%), indicating no measurable subsampling-induced bias (**Supplemental Table S4**). Our results suggest that ULI-HiFi can be used to accurately detect SNVs from low inputs of DNA in long sequencing reads.

We next compared SV data from each of the sequencing approaches. Accuracy was determined by comparing the results to the Genome in a Bottle (GIAB) benchmark set of SVs (v0.6). SVs were detected using Sniffles2, a long-read SV caller (Smolka et al. 2024). ULI-HiFi detected 32,825 SVs, and, when compared to the GIAB reference set, precision and recall were 89.57% and 91.72%, respectively, while precision and recall were 25.74% and 59.40% for dMDA, respectively (**Table 1**). With ULI-HiFi, we observed 8 correctable mistakes (Wenger et al. 2019) in the updated GIAB/T2T SV benchmark set that were not previously captured (Paulin et al. 2024**; Supplemental Table S5**). We also compared TR data from each of the sequencing approaches. Accuracy was determined by comparing the results to the Genome in a Bottle (GIAB) benchmark set of TRs (v1.0.1). TRs were detected using the Tandem Repeat Genotyper (TRGT) (Dolzhenko et al. 2024). We found that ULI-HiFi performed substantially better than dMDA (F1 values of 90.92% and 56.90%, respectively). Despite dMDA producing slightly longer reads with comparable Phred quality scores (**Supplemental Table S2**), it consistently underperformed ULI-HiFi across all variant classes. This discrepancy is likely attributable to allelic dropout, a known limitation of MDA-based amplification methods (Estévez-Gómez et al. 2025).

To evaluate the differences in genomic regions missed or identified by dMDA and ULI-HiFi, we compared the coverage distribution for these two sequencing methods using a standard QC plot (**Supplemental Fig. S1**). We observed that the percentage of bases covered by at least 10 reads were 80.96% and 69.43% for ULI and dMDA, respectively. These results suggest that ULI provides more consistent and higher coverage across the genome compared to dMDA.

To investigate the relationship between read length and quality for ULI-HiFi, we compared the read length and the predicted accuracy of HiFi reads. The predicted accuracy of the circular consensus sequencing (CCS) reads had a Phred quality score of 40 (∼99.99%) and mean read length of ∼9,000 base pairs (**Fig. 2A**). The relationship between predicted read accuracy and number of subread passes was also investigated. We observed that eight passes were required to reach Q30 (Phred quality score 30; 99.9% predicted accuracy) and 15 passes were required to reach Q40 (99.99%; **Fig. 2B** and **Supplemental Fig. S2**).

**Figure 2.**
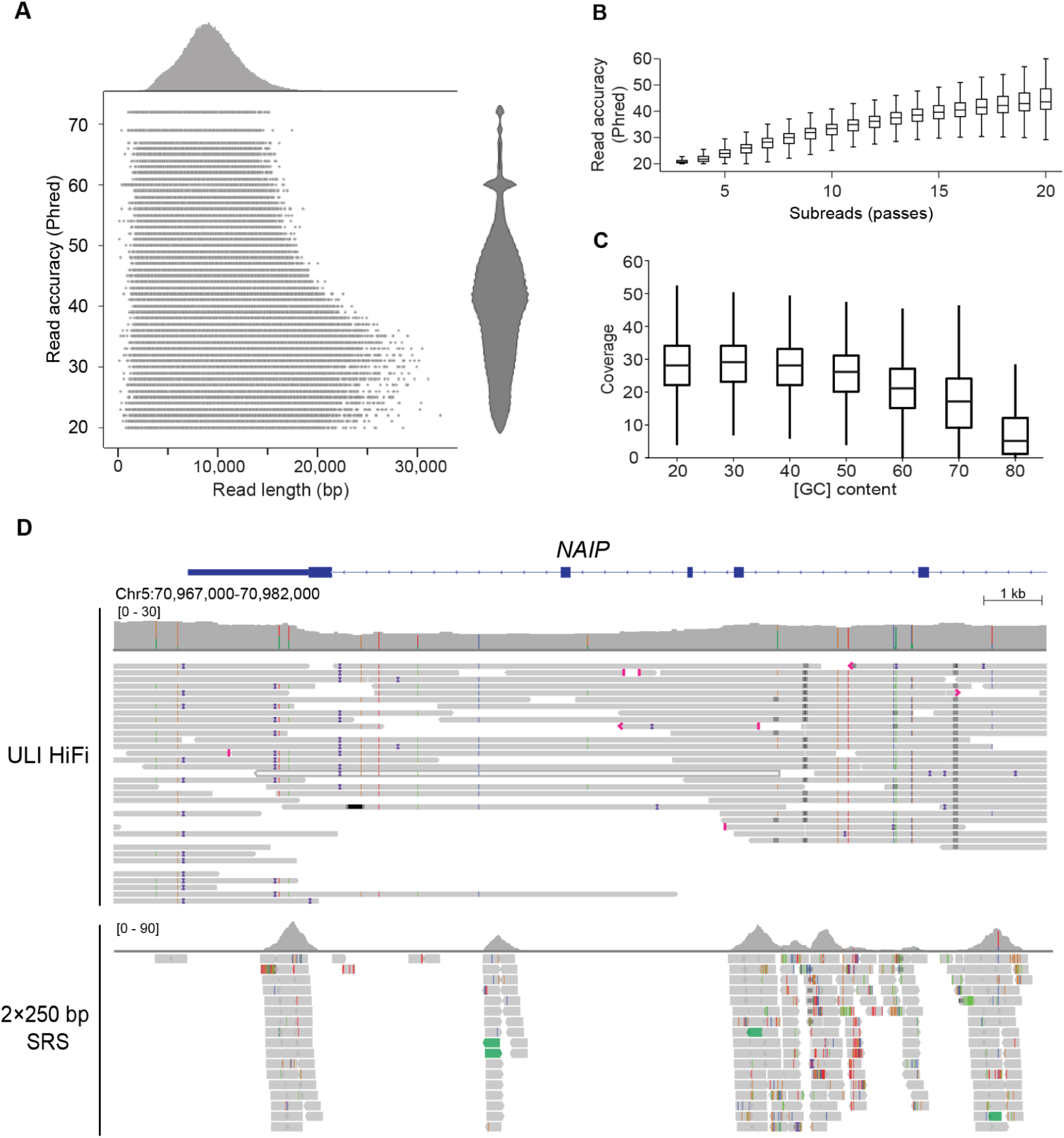
Sequencing NA24385 with ULI-HiFi reads. **A)** Sequencing read length (bp) and predicted accuracy (Phred quality score) of ULI-HiFi reads. **B)** Accuracy of reads with different numbers of passes, predicted by HiFi CCS software.**C)** Coverage distribution across genomic regions with varying GC content, measured in 500 bp windows. **D)** Coverage of the *NAIP* gene in NA24385 with 2×250 bp SRS (short-read sequencing) reads and ULI-HiFi reads. SRS reads were obtained from GIAB Illumina 2×250.

We next analyzed the distribution of reads across the genome. We divided the genome into 500 bp fixed-width windows, binned them by GC content in 10% intervals, and calculated the sequencing depth for each bin. We found that an average of 25× sequencing depth was maintained across a range of windows of varying GC content (20% to 50%), before it decreased to approximately 18× at 70% GC, showing that ULI-HiFi sequencing provides relatively uniform coverage across a broad GC content range (**Fig. 2C** and **Supplemental Fig. S3**). The ULI method allowed us to sequence several medically relevant genes that are difficult to map with SRS, such as *NAIP* (Mandelker et al. 2016; Wagner et al. 2022; English et al. 2024). ULI**-**HiFi reads revealed major portions of the gene previously unmappable with SRS (**Fig. 2D, Supplemental Table S6**). These results demonstrate ULI-HiFi’s reliability across various genomic regions and provide confidence for its use in many applications where short-read sequencing may not be sufficient.

Since repetitive regions of the genome can, in some contexts, be difficult to amplify (Ross et al. 2013), we leveraged our dataset to examine the fidelity of the ULI approach to genotype TRs. The GIAB HG002 TR v1.0.1 benchmark (English et al. 2024) was used to compare genotyping performance at 1,638,105 TR loci for the Standard HiFi and ULI samples. Overall, the two sequencing methods show comparable concordance to the reference set, with just a 3.2% decrease in perfect matches from Standard to ULI (93.6% perfect match versus 90.4% for Standard and ULI, respectively) (**Fig. 3A**). If we allowed for a single motif mismatch (Dolzhenko et al. 2024), accuracy improved to 99.7% and 98.9% for Standard and ULI respectively, indicating that most discrepancies were very small (**Fig. 3A**). When we investigated the 1% or less discordant loci (off-by-two or more motif units), we found that most of these loci (54% and 37.5% for Standard and ULI, respectively) were accurate to less than 10 motif units (**Fig. 3A**). Next, we investigated motif length. We found that ULI faithfully genotyped both short tandem repeats (STRs, motifs of 2–6 base pairs) and variable-number tandem repeats (VNTRs, motifs of ≥7 base pairs) at a level on par with Standard HiFi (**Fig. 3B**). Both methods have high levels of allele agreement up to around 500 bp where performance decreases, with VNTRs and Standard sequencing both maintaining genotyping accuracy slightly better compared to STRs and ULI (**Fig. 3B**). Additionally, both Standard and ULI reliably genotyped TRs over a broad range of GC content (99% and 97% for Standard and ULI, respectively, across repetitive regions with GC content 0–80%, (**Fig. 3C**). This range (0–80% GC) represents 99.5% of all regions in the TR catalog (1,630,318 / 1,638,105), indicating that ULI is sufficient to genotype the vast majority of TRs in the genome to within one repeat unit. At the remaining <0.5% of regions containing 80–100% GC content, both methods decrease in performance, and Standard performed better (e.g., 91% versus 70% accuracy in Standard and ULI, respectively, in 90–100% GC content regions, allowing for off-by-one motifs, **Fig. 3C**). We next studied loci associated with pathogenic repeat expansions. The ULI method was also able to genotype TRs at known pathogenic genes, performing equally well as Standard HiFi (≥95% agreement) for all but 6 of these loci (51/57, 89.5%) (**Fig. 3D**). For five of the loci, ULI failed to report a genotype due to a lack of reads spanning the locus. These results demonstrate that despite the amplification step, ULI maintains high fidelity for TR genotyping across diverse genomic contexts.

**Figure 3.**
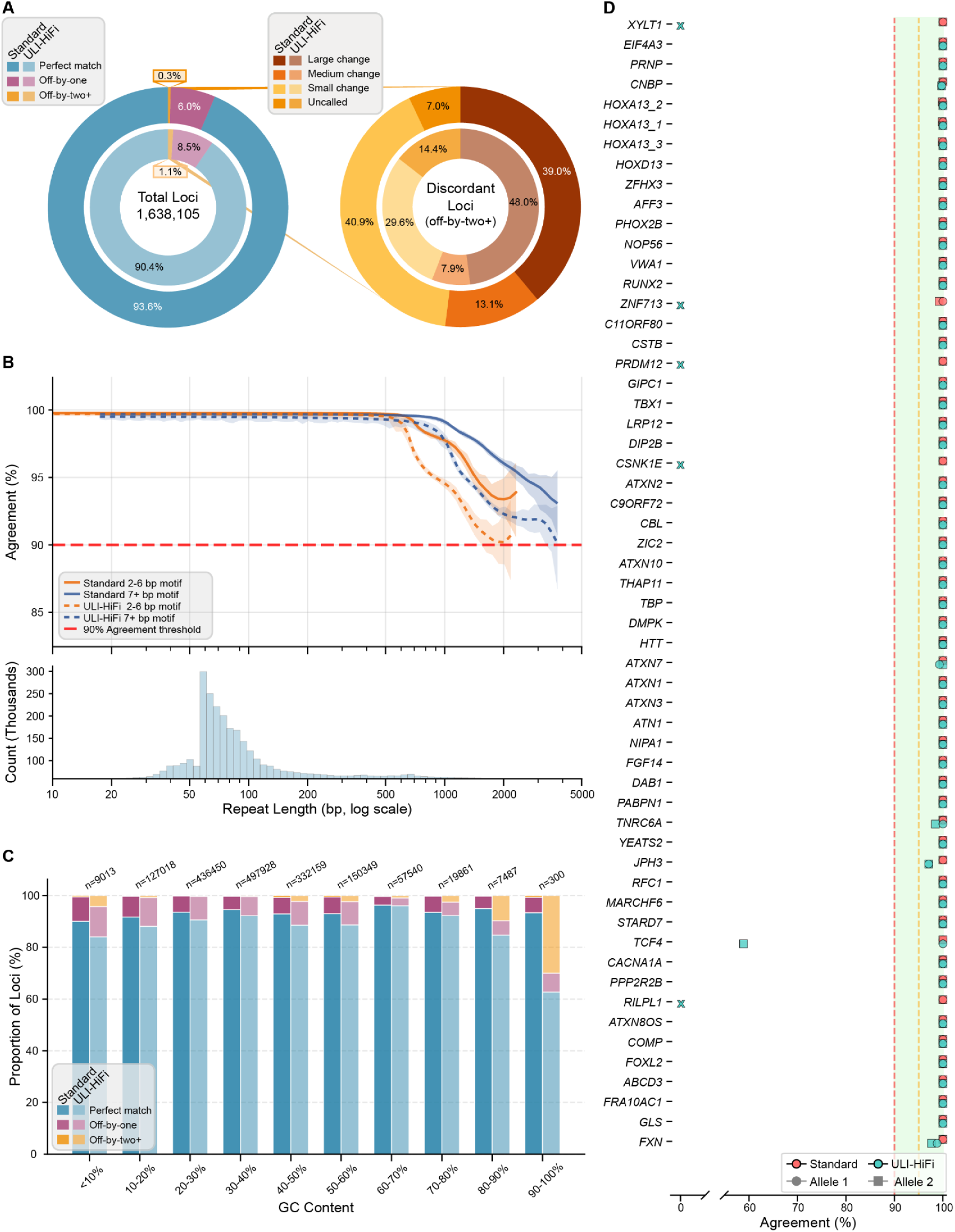
Benchmarking tandem repeat genotyping accuracy of Standard HiFi and ULI-HiFi sequencing (GIAB TR benchmark v1.0.1). **A)** Left: Concordance of genotype calls across 1,638,105 tandem repeats comparing Standard HiFi to GIAB (outer ring) and ULI-HiFi to GIAB (inner ring). Right: Classification of discordant loci by magnitude of difference (large: ≥10 motifs, medium: 5–9 motifs, small: 2–4 motifs). **B)** Top: LOWESS-smoothed genotype agreement curves for Standard HiFi (solid lines) and ULI-HiFi (dashed lines) versus GIAB, stratified by STRs (2–6 bp motifs) and VNTRs (≥7 bp motifs), with 95% confidence intervals. Bottom: Distribution of tandem repeat loci included in this analysis, binned by tract length. **C)** Genotype concordance stratified by repeat GC content for Standard HiFi (dark bars) and ULI-HiFi (light bars) compared to GIAB. **D)** Performance at medically relevant pathogenic repeat loci. Bars indicate allele-specific agreement between Standard HiFi (red) or ULI-HiFi (teal) and GIAB reference genotypes. X marks indicate failure to genotype the TR due to insufficient coverage.

We next applied this method to clinical samples. We began with a saliva sample from a healthy male donor (**Fig 4A**). This saliva sample showed similar mean read length and excellent read accuracy (Phred quality, **Fig. 4B**), as well as comparable coverage across windows of varying GC-content (**Fig. 4C**) when compared to reads generated for NA24385 genomic DNA. From this single saliva sample, we identified several high-confidence structural variants, including a 320 bp insertion in the myelin basic protein gene (*MBP*) and a 270 bp deletion in the protein tyrosine phosphatase receptor type G gene (*PTPRG*), supported by 11 and 18 reads, respectively, showing clear breakpoints (**Figs. 4D-E**). This successful application to saliva—a non-invasive, easily collected clinical sample—further demonstrates that ULI-HiFi sequencing can provide comprehensive coverage of difficult-to-sequence genomic regions from nanograms of human genomic DNA, expanding the potential applications for precision medicine approaches.

**Figure 4.**
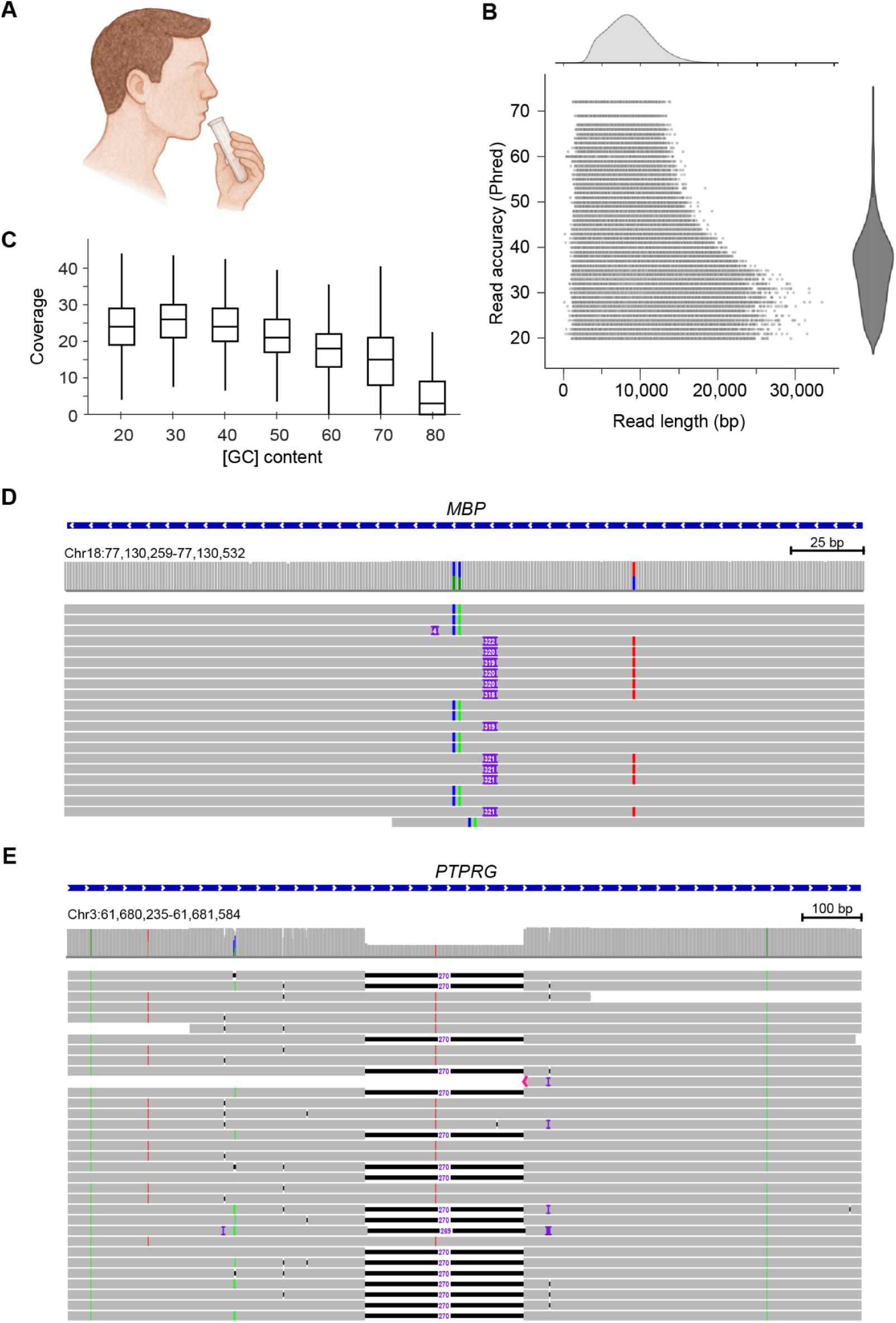
Analysis of a saliva sample with ULI-HiFi. **A)** Saliva collection from healthy adult male donor. **B)** Sequencing read length (bp) and predicted accuracy (Phred quality score) of ULI-HiFi reads. **C)** Coverage distribution of ULI-HiFi reads across genomic regions with varying [GC] content, measured in 500 bp windows. **D)** Structural variant insertion in an intron of *MBP*. **E)** Structural variant deletion in an intron of *PTPRG*.

### ULI-HiFi sequencing of a patient with familial adenomatous polyposis (FAP)

Because ULI-HiFi data is compatible with low inputs of DNA, we reasoned that it may serve as a powerful tool to analyze clinical samples with limited amounts of DNA. We applied ULI-HiFi sequencing to a patient with familial adenomatous polyposis (FAP), a hereditary form of colon cancer (**Fig. 5A**) (Becker et al. 2022). Approximately 85% of patients with FAP inherit a germline mutation in *APC*, a key tumor suppressor gene (Polakis 1997). FAP manifests as hundreds to thousands of polyps throughout the colon by the time affected individuals reach 30-45 years of age. Polyps may emerge at different time points, leading to a spectrum of malignancy across different polyps throughout the colon, some of which progress to adenocarcinoma. We analyzed a normal, polyp, and adenocarcinoma (AdCa) sample from the same patient, all with very similar sequencing depth (17 to 20×).

**Figure 5.**
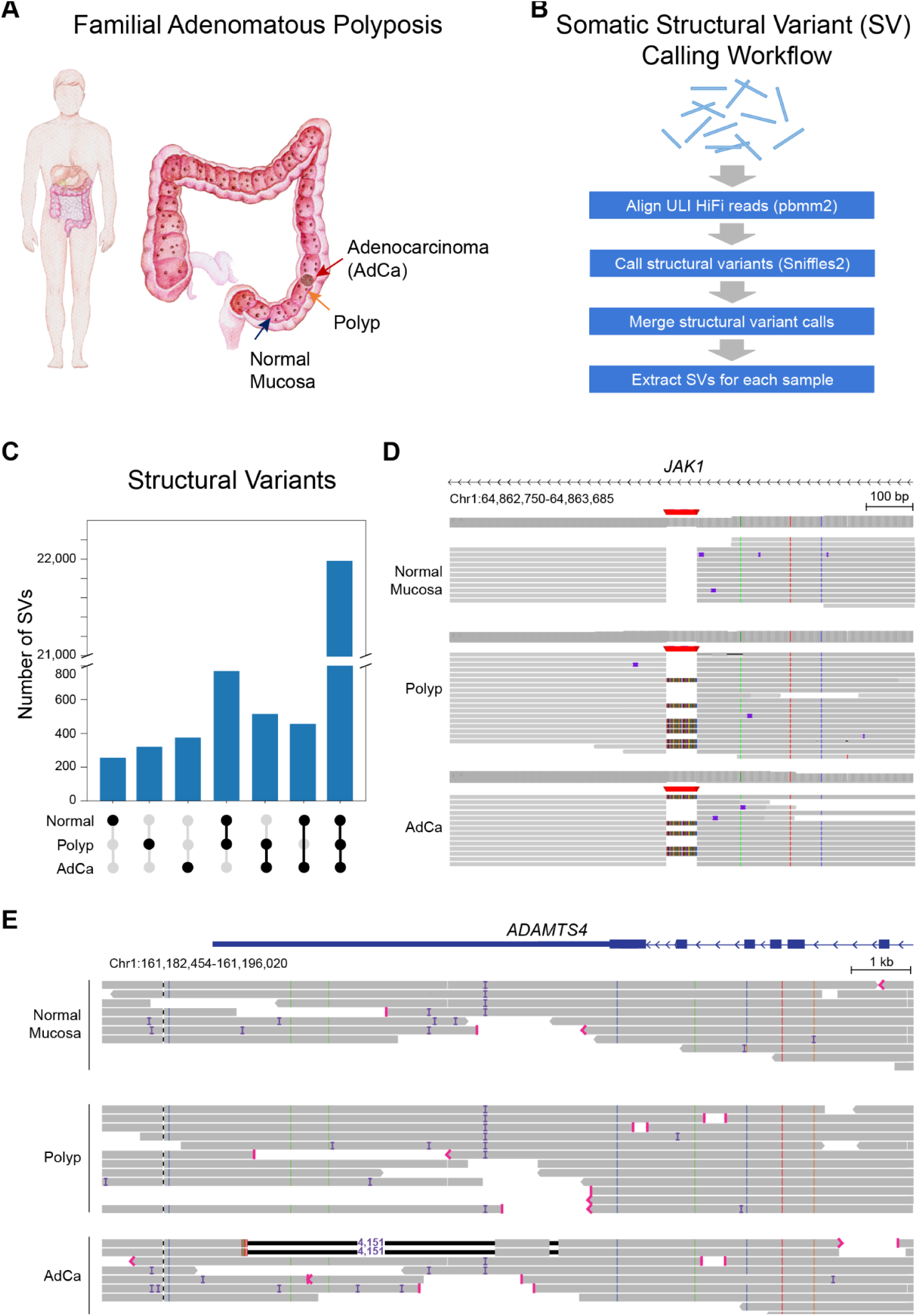
Analysis of a patient with familial adenomatous polyposis (FAP). **A)** Colon from a patient with familial adenomatous polyposis (FAP). This illustration shows the location of the normal tissue sampled, the polyps, and the adenocarcinoma (AdCa). Note, the lesions are on the lumen side of the colon. **B)** Scheme to describe the somatic structural variant (SV) detection workflow. **C)** Overlap of structural variants (SVs) identified between normal, polyp, and adenocarcinoma (AdCa) samples. Each bar represents a specific pattern of SV occurrence across tissues, with the dot matrix below indicating which tissues share those variants. The first three bars from the left show SVs unique to single tissues (normal only, polyp only, AdCa only), the middle three bars represent SVs found in exactly two tissue types, and the rightmost bar shows SVs common to all three tissues. **D)** Example of a SV identified in the intron of *JAK1*. **E)** Mosaic deletion detected in adenocarcinoma (supported by 2 reads), located in the 3′ UTR of *ADAMTS4*.

Given our ability to detect SNVs with ULI-HiFi, we also analyzed SNVs in polyp and adenocarcinoma samples. We first looked to see whether we could observe the likely pathogenic *APC* mutation by sequencing adjacent normal colon tissue in the FAP patient. Indeed, we observed a germline mutation in a splice donor site of *APC* that was previously identified as likely pathogenic (Aretz et al. 2004**; Supplemental Fig. S4**).

Next, we examined SVs in this patient with FAP. One of the major advantages of long-read DNA sequencing, including ULI-HiFi sequencing, is its ability to identify large SVs that are frequently missed by short-read DNA sequencing (Logsdon et al. 2020; Marwaha et al. 2022). We used Sniffles v2.2 to call somatic SVs in normal, polyp, and adenocarcinoma samples with reference genome hs37d5 (**Fig. 5B** and **Methods**), and extracted unique calls by filtering for SVs found only in adenocarcinoma samples or only in polyp samples. We manually inspected 100 randomly selected SVs from this callset and identified supporting reads for 84 of the 100 (84%).

We identified 21,197 SVs in the normal mucosa, the pre-cancerous polyp, and the adenocarcinoma samples (**Fig. 5C**). Notably, the number of unique SVs identified increased from normal mucosa to polyp to adenocarcinoma (256, 320, and 375, respectively), suggesting that large-scale SVs are associated with FAP polyp development and progression (**Methods**). Importantly, this observation was independent of sequencing read depth. One SV of particular interest was identified in the intron of *JAK1*, a known oncogene (Jeong et al. 2008). This SV, an insertion of 66 base pairs, was observed in polyp and adenocarcinoma samples, but not in the normal mucosa (**Fig. 5D**). Additionally, a mosaic deletion of ∼4,100 base pairs present only in the adenocarcinoma sample was identified in the 3′ UTR of *ADAMTS4*, supported by two sequencing reads (**Fig. 5E**). This gene, *ADAMTS4*, is often upregulated in cancer (Shang et al. 2020). Overall, ULI-HiFi revealed thousands of SVs across a range of tissue types.

Based on the promising results with SNVs and SVs and from our detailed benchmarking of ULI, we proceeded to analyze tandem repeat (TR) DNA sequences. We proceeded to genotype the normal, polyp, and adenocarcinoma samples across the catalog of 937,122 TRs (Dolzhenko et al. 2024). We observed 31,781 changes in genotyped TR lengths between normal and polyp and 36,335 changes between normal and adenocarcinoma (**Fig. 6A**). Among these TR variations, we observed a heterozygous, mosaic TR expansion of an AC motif in the 5′ UTR of *LIMD1*, which caught our attention (**Fig. 6B**). Repeat length progressively expanded from 57 copies in the normal sample to 61 and 74 copies in polyp, and adenocarcinoma samples, respectively. Manual inspection of the sequencing reads revealed 6 and 2 supporting reads for the polyp and adenocarcinoma genotypes, respectively (**Fig. 6B**). Since the original tissue sample was depleted during initial analyses, we pursued an alternative approach to characterize this repeat expansion in an independent dataset. To do this, we leveraged ExpansionHunter to examine short-read DNA sequencing data from the Pan-Cancer Analysis of Whole Genomes (PCAWG) dataset (Dolzhenko et al. 2019). This technique has successfully been applied to accurately genotype both known and novel repeat expansions (Dolzhenko et al. 2020; Fazal et al. 2020; Watanabe et al. 2022; Méreaux et al. 2023). We analyzed 2,658 paired short-read whole genome sequencing samples from the PCAWG dataset, covering 29 distinct cancer types. We identified 49 tumor samples with expansions of the AC repeat in the 5′ UTR of *LIMD1* across these samples (**Supplemental Table S7**). The recurrence of this specific expansion across multiple independent tumor samples corroborates our initial findings and prompted us to investigate its functional significance.

**Figure 6.**
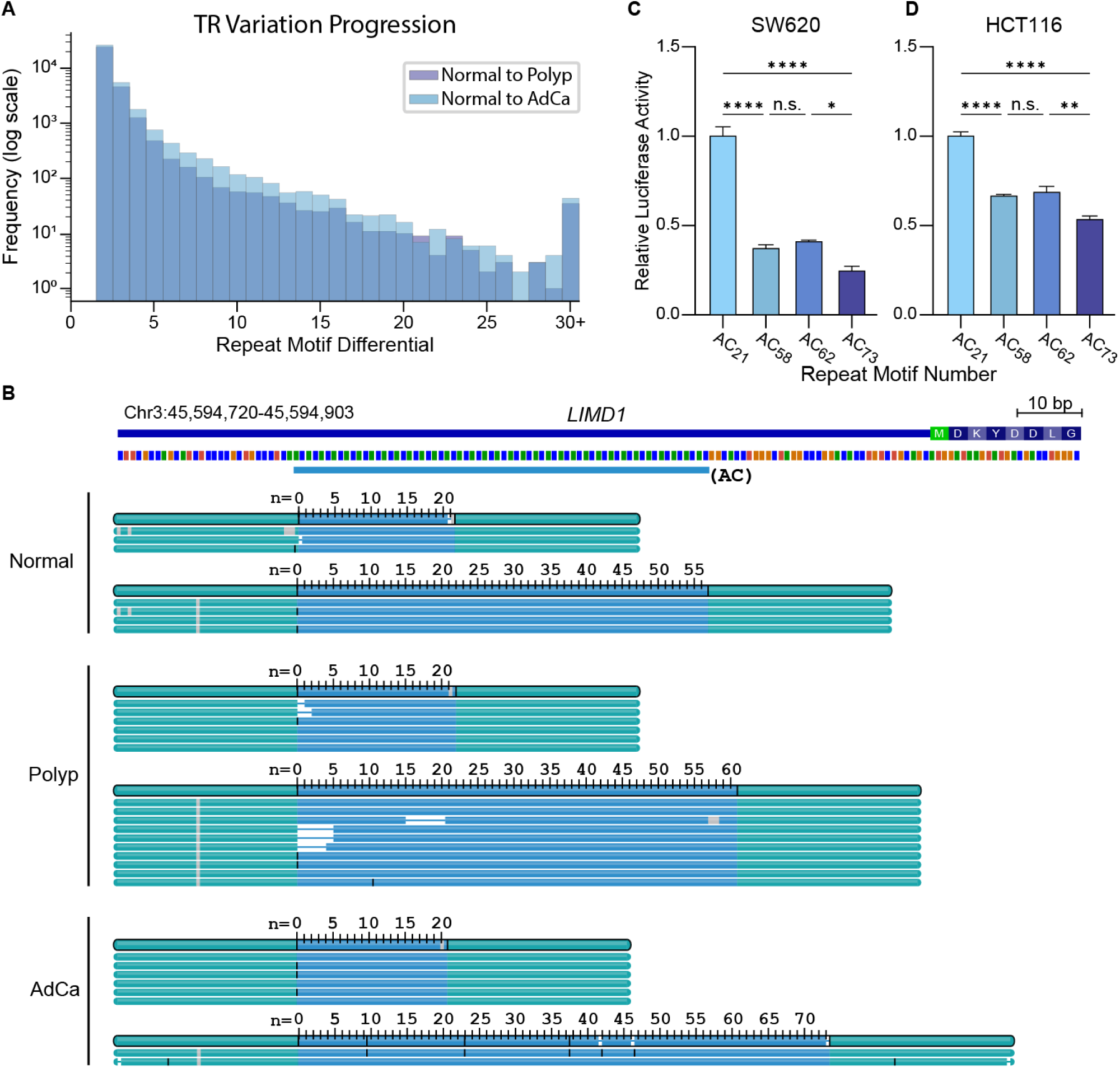
ULI-HiFi reads genotype tandem repeats. **A)** TR variation between normal and polyp (indigo) and between normal and adenocarcinoma (AdCa, light blue). Regions with a repeat motif differential of 1 are excluded. **B)** TRGT visualization of *LIMD1* mosaic TR expansion from normal to polyp to AdCa tissue. Flanking sequences are shown in green and the repeat region is shown in blue, with counts corresponding to the number of AC repeat units. Sequencing depth ranged from 7–16× for this region. **C, D)** Luciferase activity of two colorectal cancer cell lines, SW620 (C) and HCT116 (D), with nano-luciferase under control of *LIMD1* promoter and 5′ UTR harboring different AC repeat lengths, as indicated in the figure. Results are means ± SEM, normalized to the expression of the non-expanded allele (21 motif units). \**P_adj_* < 0.05, \*\**P_adj_* < 0.01, \*\*\*\**P_adj_* < 0.0001 by one-way ANOVA with post-hoc Tukey test.

Having observed the recurrence of the *LIMD1* TR expansion across multiple tumors, we next assessed its impact on gene expression through luciferase reporter assays. We engineered a series of plasmids with the *LIMD1* promoter and 5′ UTR containing AC repeats that faithfully represented those observed in patient samples (+/− 1 repeat unit accuracy despite the well-known technical challenges of repeat stability during molecular cloning) and used these constructs to drive Nano-luciferase expression. Using these engineered constructs, we observed a significant decrease in luciferase expression with increasing repeat length (**Figs. 6C, D**). This decrease was observed in both an *APC* wild-type colorectal cancer cell line (HCT116) and an *APC*-mutant CRC cell line (SW620). These results, along with the established role of *LIMD1* as a tumor suppressor gene across multiple cancer types (Sharp et al. 2004; Zhang et al. 2018; Chakraborty et al. 2018), suggest further studies investigating this tandem repeat expansion are warranted.

## Discussion

Here, we benchmarked two methods to perform long-read sequencing from an ultra-low input (ULI) amount of DNA sample. We applied the protocol to sequence the reference human sample, NA24385 (HG002), comparing different amplification methods (dMDA and ULI-HiFi). Our findings reveal (i) when comparing two methods of DNA amplification, the ULI-HiFi method afforded better precision and recall rates for small variants, particularly SNVs; (ii) ULI-HiFi detected SVs and TRs with relatively high precision and recall (e.g., 98.9% accuracy for TRs within ±1 motif), and significantly outperformed dMDA; (iii) ULI-HiFi sequencing enabled access to clinical samples with low sample inputs, as demonstrated by the sequencing of a saliva sample from a healthy participant, and a normal, polyp, and adenocarcinoma from a patient with familial adenomatous polyposis (FAP); and (iv) ULI-HiFi enabled the discovery of SVs in both known oncogenes and tumor suppressor genes and revealed a TR expansion in *LIMD1*, a gene implicated in tumorigenesis.

In the past, long-read sequencing technologies have been limited to the study of samples with several micrograms of DNA, such as cell lines and whole blood. By contrast, ULI-HiFi requires just 10 nanograms of DNA input. This small sample requirement allows for the analysis of precious clinical samples, which we demonstrated here. Note added during revision: PacBio announced SPRQ chemistry reducing standard HiFi input requirements to 500 ng; however, ULI-HiFi’s 50-fold lower requirement (10 ng) remains advantageous for precious clinical samples. Though our cancer analysis consists of three samples from one patient, it serves to substantiate our claim that the ULI-HiFi method can be used for clinically relevant analysis of patient DNA, alongside the thorough benchmarking performed here. With long-read sequencing, researchers can investigate many additional regions of the genome that are invisible to short-read sequencing due to an inability to map short reads uniquely in the genome. ULI-HiFi’s ability to resolve genomic regions with complex repeat structures and high GC content enable its coverage of medically relevant regions of the genome and identification of variants otherwise missed by traditional short-read sequencing. As evidenced by the necessity of long-read technology to fill the gaps in the telomere-to-telomere sequencing of the CHM13 genome, short-read sequencing leads to large gaps in our understanding of the genome (Miga et al. 2020). The ULI-HiFi method shows good sensitivity and precision against the GIAB reference set of SVs and TRs. Comparing SVs between normal mucosa, pre-cancerous polyps, and an adenocarcinoma, our method identified several SVs found exclusively in the adenocarcinoma. Several cancers lack a mutation in a known cancer driver gene (Macintyre et al. 2016), and this initial study demonstrates that the ULI-HiFi approach may enable a deeper understanding of the genetic alterations in these cancer types and may provide new insights into oncogenic transformation.

Historically, PCR-based genotyping of TRs has been notoriously challenging due to amplification bias (e.g., preferential amplification of shorter alleles or failure in GC-rich regions and regions that form secondary structures) (Aird et al. 2011). Some repeats (e.g., the GAA expansion in the *FXN* gene, the cause of Friedreich ataxia) can be genotyped with specialized PCR assays, while other repeats have been deemed “unsequencable” (Loomis et al. 2013). These long-standing challenges raised the question of whether a whole-genome amplification method would corrupt TR length, leading to unfaithful genotyping. Our results provide the first comprehensive evidence that this is not the case: compared to a reference set of >1.6 million TR genotypes (English et al. 2024), we found that >90% of ULI-HiFi TR genotypes were in perfect concordance with the truth set, and 98.9% matched to within a single repeat unit. This study represents the first large-scale demonstration that PCR-based amplification can faithfully genotype the vast majority of TRs to within a single repeat unit, thereby expanding the utility of amplification-based approaches for comprehensive TR analysis in clinical and research applications where input DNA is limiting.

Despite its benefits, there are some limitations to the ULI-HiFi method. First, we observed reduced accuracy of indel calls compared to the standard amplification-free HiFi method. Any amplification-based method, including ULI-HiFi, may have a propensity for introducing indel mutations, which are usually found in repetitive homopolymeric sequences of the genome. Future efforts to optimize amplification are warranted. The second limitation is the loss of DNA methylation information. One of the features of standard HiFi is the ability to detect DNA methylation at single-base resolution. PCR-amplified DNA lacks methylated cytosines, which means that methylation status is not recovered with ULI-HiFi. Additionally, when working with ultra-low quantities of clinical samples, our current coverage depth sometimes results in reported variants supported by the minimum threshold of reads, particularly in heterogeneous tissues. The threshold used here is consistent with previous successful detection of pathogenic SVs (Smolka et al. 2024; Jiang et al. 2020) and in the detection of pathogenic SVs in a clinical setting (Merker et al. 2018); this, in addition to our comprehensive benchmarking against the gold-standard reference set (Genome in a Bottle), provides confidence in our findings. However, future studies would benefit from increased sequencing depth to provide additional read support for such variants.

The ULI-HiFi approach described here can be used to uncover dark regions of the genome from a fraction of the DNA that other long-read technologies require. By requiring orders of magnitude less input DNA and maintaining 99%+ accuracy for SNV and TR variant detection, ULI-HiFi opened the door to previously impossible clinical applications—culminating in the identification of a stepwise *LIMD1* expansion with evidence supporting a functional role in a single patient’s colon. Our work also challenges the longstanding notion that many TRs cannot be faithfully amplified and genotyped. The ULI-HiFi approach will enable access to previously unexamined regions of the genome and set the stage for a better understanding of human disease.

## Methods

### Sample Preparation

NA24385 (HG002) DNA was obtained from Coriell (Coriell Institute for Medical Research, Camden, New Jersey) and used without further purification. Twenty (20) nanograms of DNA was used for the ULI-HiFi procedure, as described below. The adjacent normal tissue (49.38 mg, 30 mm), polyp (34.22 mg, 5 mm), and adenocarcinoma (34×17×10 mm) samples from patient A001 were obtained from the descending colon of a patient in their fifties with familial adenomatous polyposis during a colectomy. Several milligrams of fresh tissue were obtained and snap-frozen in liquid nitrogen. The tissue was pulverized with mortar and pestle, and DNA was extracted using Qiagen All-prep kit (cat. 80204). DNA was isolated as described elsewhere (Becker et al. 2022), yielding several micrograms of genomic DNA. However, the majority of the sample was used in prior analyses, leaving 1168, 166, and 60 nanograms of genomic DNA from normal, polyp, and adenocarcinoma, respectively, in this study. Ten (10) nanograms of each sample was used for the ULI-HiFi procedure, as described below. The quality of the genomic DNA was assessed using standard spectrophotometric measurements of 260/280 and 260/230 to ensure purity. The saliva samples were obtained from an adult male through a joint effort between PacBio and DNA Genotek.

### Sample Processing

#### Shearing, Ultra-Low Input (ULI) Amplification, and Analysis of Sample Quality

NA24385 (20 ng) and A001 (10 ng each of normal, polyp, and adenocarcinoma) genomic DNA was sheared to approximately 11 kb with a Covaris g-TUBE following manufacturer’s instructions. The results of the shearing were evaluated with a Femto Pulse system (Agilent Technologies, Santa Clara, California). For each sample, sheared DNA (10 ng) was processed with the SMRTbell gDNA sample amplification kit following manufacturer’s instructions (Catalog 101-980-000, PacBio, Menlo Park, California). Libraries were size-selected >8 kb with a Blue Pippin system (Sage Science, Beverly, Massachusetts).

To assess DNA quality prior to library preparation, we employed three measures. DNA concentration was quantified with a Qubit fluorometer with the dsDNA High Sensitivity assay kit (Thermo Fisher Scientific). DNA purity was measured by absorbance with a NanoDrop spectrophotometer; samples with A260/280 ratios near 1.8 and A260/230 ratios ≥ 2.0 were prioritized for library preparation. Fragment size distribution was analyzed by pulsed-field capillary electrophoresis with a Femto Pulse system (Agilent Technologies). Post-shearing profiles were also evaluated to confirm an appropriate fragment size distribution centered near the target insert size (∼11 kb).

#### Droplet Multiple Displacement Amplification (dMDA)

A total of 0.45 ng of genomic DNA, corresponding to approximately 14 pg of DNA per reaction, was mixed with dMDA reagents and encapsulated into single emulsion droplets by Xdrop™. In total, 32 reactions were performed to make enough amplified DNA for PacBio HiFi sequencing. Following a 16-hour incubation, the amplified DNA was isolated and subjected to T7 endonuclease I treatment to remove branching. Lastly, DNA repair was performed and Tapestation was used to estimate DNA molecule size (Madsen et al. 2020).

### HiFi Sequencing

Samples from NA24385 (Standard PacBio, dMDA) and A001 were sequenced on a PacBio Sequel II system with the Sequel II binding kit 2.0 and chemistry 2.0 (Catalog 101-789-500 and 101-820-200, PacBio, Menlo Park, California). These samples were sequenced on the SMRT Cell 8M platform, with 2 SMRT cells used for NA24385 Standard 28× HiFi, A001 adenocarcinoma samples, and A001 polyp samples, 3 SMRT cells used for A001 normal samples, and 4 SMRT cells used for dMDA to ensure sufficient coverage (coverage >15×) (**Supplemental Table S4**) (Harvey et al. 2023). The normal donor saliva sample and NA24385 (ULI) were sequenced on a PacBio Revio system following Megaruptor 3 shearing and preparation with the SMRTbell prep kit 3.0 (Catalog 102-141-700, PacBio, Menlo Park, California). These samples were sequenced on the SMRT Cell 25M platform, with 1 SMRT cell used for the saliva sample and NA24385 ULI each, since the SMRT Cell 25M platform provides greater coverage per cell than SMRT Cell 8M. When multiple SMRT cells were used for a single sample to increase coverage, a single library was prepared, and that library was run across multiple SMRT cells. Libraries were sequenced with 2-hour pre-extension and 30-hour movie time. Consensus reads (CCS reads) were generated with ccs software v.3.0.0 (https://github.com/pacificbiosciences/unanimity/) with `--minPasses 3 --minPredictedAccuracy 0.99 --maxLength 21000`. Each ULI-HiFi movie was preprocessed by trimming adapters using lima and performing deduplication using pbmarkdup.

### Data Processing

2×250 NA24385 SRS reads were obtained from GIAB (https://ftp.ncbi.nlm.nih.gov/ReferenceSamples/giab/data/AshkenazimTrio/HG002_NA24385_son/NIST_Illumina_2x250bps/).

PacBio Standard HiFi NA24385 reads from the PacBio Sequel II system were obtained from GIAB (https://ftp.ncbi.nlm.nih.gov/ReferenceSamples/giab/data/AshkenazimTrio/HG002_NA24385_son/PacBio_CCS_15kb_20kb_chemistry2/reads/).

PacBio Standard HiFi NA24385 reads from the PacBio Revio system were obtained from GIAB (https://ftp-trace.ncbi.nlm.nih.gov/ReferenceSamples/giab/data/AshkenazimTrio/HG002_NA24385_son/PacBio_HiFi-Revio_20231031/).

NA24385 CCS reads for ULI, dMDA, and Standard HiFi were processed through the PacBio Human WGS Snakemake Workflow (https://github.com/PacificBiosciences/pb-human-wgs-workflow-snakemake) on an AWS EC2 instance to align them to GRCh38 using pbmm2 v1.10.0 and call small variants, except for the analysis of SVs in NA24385, for which we separately aligned the data to hs37d5. Where listed, samples were downsampled with `samtools` `view` `-s` using SAMtools version 1.19 (Danecek et al. 2021) to match read depth across datasets. Resulting metrics were averaged across the three datasets.

### Variant Calling

Small variants (SNVs and indels) were called with DeepVariant v1.4.0 with the PacBio model as contained in the PacBio Human WGS Snakemake Workflow. (Poplin et al. 2018).

#### NA24385

SVs for NA24385 were called with Sniffles v2.2 with default parameters and pbsv 2.9.0 (https://github.com/PacificBiosciences/pbsv) with parameters -A 2 -O 2 -S 0 -P 30 -t INS,DEL.

TRs for NA24385 were called with Tandem Repeat Genotyper (TRGT) v.1.1.1 with default parameters. The output VCF files were sorted and indexed with SAMtools v1.18.

#### Patient A001 normal, polyp, and adenocarcinoma

SVs for A001 were called with Sniffles v2.2 using default parameters along with the `--snf` tag. To compare SV calls across the adenocarcinoma, polyp, and normal samples, Sniffles2 VCF SV files were processed through a somatic SV calling workflow, with a threshold of 2 supporting reads for SV detection, meaning a variant had to be present in 11% of the reads to be considered a somatic variant. The SNF files were merged and SVs for the normal, polyp, and adenocarcinoma samples were extracted using the `SUPP_VEC` function for BCFtools (Danecek et al. 2021). In-depth analysis of specific SVs unique to the adenocarcinoma sample was conducted by visualizing loci with Integrative Genomics Viewer (IGV) (Robinson et al. 2011).

### Tandem Repeat (TR) analysis of NA24385; normal, polyp, and adenocarcinoma from patient A001

#### NA24385

We genotyped across all NA24385 samples listed in Table 1 using the Tandem Repeat Genotyper (TRGT) version 1.1.1 with default parameters. The genotyping was performed against the adotto v1.2 TR catalog (English et al. 2024), which consists of 1,784,804 TR loci. The output VCF files were sorted and indexed with SAMtools v1.18.

We employed TRGT v1.1.1 with default parameters on ULI-HiFi and Standard HiFi (no amplification) reads from NA24385 to determine if genome amplification altered the results of TR genotyping.

#### Patient A001 normal, polyp, and adenocarcinoma

TRs were genotyped in the normal, polyp, and adenocarcinoma sample of patient A001 to identify changes present across the three samples (TRGT v1.1.1 with default parameters) using the catalog of 937,122 TRs. A TR expansion was defined to be an increase in motif count greater than one (e.g., (TA)2 to (TA)4). Tandem Repeat Visualizer (TRVZ) was used to visualize tandem repeat expansions.

To genotype the STR expansions originally identified in A001 ULI-HiFi data by TRGT using an orthogonal approach, we ran ExpansionHunter v5.0.0 on A001 short-read BAM files with default parameters and its accompanying repeat catalog. The resulting BAMlet files, containing aligned reads relevant to the detected repeat expansions, were sorted and indexed with SAMtools v1.18 and visualized with REViewer v0.2.7 (Dolzhenko et al. 2022).

### Benchmarking of NA24385

Small variant callsets were benchmarked against GIAB HG002 set v4.2.1 with hap.py v3.12.1 (https://github.com/Illumina/hap.py) with parameters `--pass-only -r ./human_GRCh38_no_alt_analysis_set.fasta -f ./HG002_GRCh38_1_22_v4.2.1_benchmark_noinconsistent.bed --engine=vcfeval`. Only PASS calls were included.

SVs were benchmarked against GIAB HG002 set v0.6 (ftp://ftp-trace.ncbi.nlm.nih.gov/giab/ftp/data/AshkenazimTrio/analysis/NIST_SVs_Integration_v0.6) with Truvari v4.1.0 (https://github.com/spiralgenetics/truvari). Truvari was run with parameters `--includebed HG002_SVs_Tier1_v0.6.bed --passonly -r 1000 -p 0.0 --sizemin 50 --includebed HG002_SVs_Tier1_v0.6.bed.` TRs were benchmarked against the 1,638,105 tier 1 loci in the GIAB HG002 set v1.0 loci with Truvari v4.3.0. To benchmark variants, Truvari bench was run with parameters `--sizemin 50 –pick ac`. To harmonize phased variants and find more matches, Truvari refine was run with the `-w` option and otherwise default parameters.

For in depth comparison of TR genotyping performance, the Tier 1 loci were selected from TRGT VCFs of the GIAB HG002 v1.0.1 benchmark, ULI 27.5×, and Standard samples. Concordance was defined as the proportion of alleles in the bin that agreed with the benchmark alleles following these rules: match = x < 0.5 motif difference, off-by-one = 0.5 ≤ x < 1.5, and discordant = 1.5 ≤ x motif difference. Allele percent agreement was defined as follows, with alleles paired to maximize mean agreement across call sets:

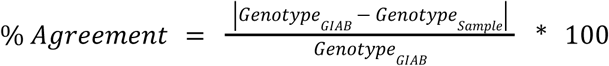

### Analysis of NA24385

The average read length and read quality were calculated using the read length and quality script contained in the PacBio Human WGS Snakemake Workflow. Gene visualization was performed using Integrative Genomics Viewer (IGV) for HG002 2×250 bp SRS reads and ULI-HiFi reads.

To measure coverage distribution across genomic regions with varying GC content, alignment files from 2 SMRTcells were merged and sorted using SAMtools v1.13. To measure coverage per single-base resolution, `samtools` `mpileup` with default perameters was used. The final GC content (%) was computed every 500bp window with no overlaps of the GRCh38 reference genome. Average coverage was also computed for approximately 5.5M windows.

To compare the coverage distribution between ULI and dMDA, the percentage of bases covered by at least 10 reads for ULI and dMDA were calculated in 1 kb bins. To account for coverage correlation in adjacent regions and reduce covariance for statistical analysis, coverage values were binned in 10 kb bins using the command `bedtools` `makewindows` from BEDTools v2.31.1 with default parameters (Quinlan and Hall 2010). `bedtools` `map` with default parameters was used to classify bins as covered based on whether their median coverage exceeded 10 reads. Proportions of covered bins for each sample were compared using a two-proportion Z-test based on the proportions (p1 = 0.84502, p2 = 0.778597) and total number of bins (n1 = 321184, n2 = 321184) for ULI and dMDA samples, respectively.

To assess the genomic coverage differences between ULI and dMDA across medically relevant genes, mosdepth coverage files output from the PacBio Human WGS Snakemake workflow representing binned coverage per base pair were used. mosdepth v0.3.9 was run separately on NA24385 SRS 2×250 reads with parameters `--by 1` to act as a control (Pedersen and Quinlan 2018). The coverage information contained in the output files was normalized for each base using a normalization factor calculated as the actual sequencing coverage divided by the desired sequencing coverage, which was set as 15.5× representing the sequencing method with least coverage (dMDA). The `bedtools` `map` command with default parameters was used to map the normalized coverage values to the medically relevant gene sets.

All analysis was performed on the sample of NA24385 DNA that was sheared with 20 ng except for accuracy of subreads, which began shearing with a larger quantity of DNA. However, still only 10 nanograms were used in the libraries for sequencing.

### TR analysis in pan-cancer analysis of whole genomes (PCAWG) dataset

To genotype the AC expansion in *LIMD1* in a larger cohort of patient samples, we analyzed 2,658 paired short-read Illumina sequencing samples from the Pan-Cancer Analysis of Whole Genomes (PCAWG) dataset. Tandem repeat genotypes were assessed using ExpansionHunter v5.0.0 as described above (Dolzhenko et al. 2019). ExpansionCooker (https://github.com/lufinot/expansioncooker) was employed to identify read-supported changes between each paired tumor-normal sample. ExpansionCooker filters out alleles that lack sufficient read support on either side of the repeat as described below, ensuring only well-supported calls are considered. ExpansionCooker requires a minimum of two reads spanning the entire repeat to be counted, with total repeat coverage set to a minimum of 8. The total coverage is calculated by treating each spanning read as 1 and each partially overlapping read (not fully covering the repeat) as 1/4. Both types of reads contribute to the total coverage. Only paired samples showing a difference of 4 or more base pairs between normal and cancer samples (equivalent to two repeat motifs) were retained, to account for potential off-by-one errors from sequencing or alignment artifacts.

The resulting BAMlet files were sorted and indexed using SAMtools v1.18. Read-level support and allele-specific coverage were visualized and evaluated using REViewer v0.2.7. Expansions were categorized by the quality of read support. Tier 1 included clear expansions with strong read coverage, while Tier 2 comprised cases with ambiguous alignments or potential allele dropout. This analysis identified 74 instances of *LIMD1* AC repeat expansions, with 49 classified as Tier 1 and 25 as Tier 2 (**Supplemental Table S7**).

### Luciferase Assay

The molecular consequences of the AC repeat in *LIMD1* were examined using the Nano-Glo Dual Luciferase Reporter Assay system (Promega, N1620), using pGL4.53 (Promega, E5011) as the control plasmid and pNL1.1 (Promega, N1001) as the experimental plasmid. The NanoLuc (Nluc) locus of the pNL1.1 plasmids were altered to be under the control of the *LIMD1* promoter and 5′ UTR, which harbored AC repeat numbers based on the analysis of the FAP patient DNA (21, 57, 61, or 74 AC repeats). To insert AC repeats of defined lengths into pNL1.1 plasmids, two rounds of InFusion cloning (Takara Bio, 638946) were performed. In the first round of cloning, HG002 DNA from the *LIMD1* promoter and 5′ UTR was inserted into a primer-linearized pNL1.1 plasmid. These clones were verified with Sanger DNA sequencing (Azenta Life Sciences GeneWiz). To insert the variable lengths of AC repeats, the vectors with HG002 *LIMD1* promoter and 5′ UTR upstream of Nluc were linearized and PCR-amplified, with the native repeat region excluded. Four oligonucleotides containing the desired repeat lengths (21, 57, 61, and 74 AC repeats) (Integrated DNA Technologies) were inserted into the new linearized vectors with the second round of InFusion cloning (Takara Bio, 638946). Final plasmid sequences were verified with Sanger sequencing upon creation (Azenta Life Sciences GeneWiz) and Oxford Nanopore sequencing (Plasmidsaurus). We were able to successfully clone plasmids with 21, 58, 62, and 73 AC repeats, which are all within one repeat motif of the original genotypes.

Colorectal cancer cells were seeded in quintuplicate into white 96-well plates (Nunc, 136101) at 25% confluency. After 24 hours, the cells were co-transfected with pGL4.53 and one of four pNL1.1 plasmids with varying repeat lengths as reported. Transfection was performed with Lipofectamine 3000 (Invitrogen, L3000001) according to the manufacturer’s protocol. The Nano-Glo Dual Luciferase Reporter Assay was performed 24 hours post-transfection following the manufacturer’s protocol, and luciferase activity was measured with a CLARIOstar Plus microplate reader (BMG Labtech). Luminescence values are reported as mean normalized RLU values with pGL4.53 as the internal control ± SEM.

### Cell culture

SW620 cells were maintained in DMEM 4.5 g/L (Gibco, 11965-092) + 10% FBS (Avantor, 89510-1186) + penicillin/streptomycin 250 U/mL/250 μg/mL (Gibco, 15070-063). HCT-116 cells were maintained in McCoy’s 5A medium (Gibco, 16600-082) + 10% FBS + penicillin/streptomycin 250 U/mL/250 μg/mL. All cells were incubated at 37°C, in a humidified atmosphere of 5% CO_2_. Cell growth and density were monitored using phase contrast microscopy, and cell viability was quantified with trypan blue exclusion with a Countess 3 (Thermo Fisher Scientific). Cells were passaged with 1× TrypLE (Gibco, 12604-021) followed by centrifugation at 300g for 5 minutes and resuspension in the appropriate fresh media.

### Statement on Software Use

Adobe Illustrator was used to assemble all of the figures in the manuscript. Sonnet 3.7 was minimally used to improve the flow of some sentences, always with a human reviewing and modifying the suggestions. A combination of gpt-image-1 and Adobe Photoshop was used to generate the scheme in Fig. 4A. The other schemes (Figures 1 and 5A) were generated by hand. There was no AI use in the analysis or interpretation of data.

## Data Access

All raw and processed sequencing data from HG002 generated in this study have been submitted to the NCBI BioProject database (https://www.ncbi.nlm.nih.gov/bioproject/) under accession number PRJNA1005794. Sequencing data and called structural variants from A001 have been submitted to NCBI dbGaP (https://dbgap.ncbi.nlm.nih.gov/) under accession number phs002371. PacBio ULI-HiFi data from the saliva sample were deposited in EGA (https://ega-archive.org/) under accession EGAS50000001666.

## Acknowledgments

This work was supported by NIH U2CCA233311 and 1U2CCA233311 to M.P.S., NIH R00HG011467, CPRIT RR23004, and a PacBio SMRT grant to G.S.E. G.S.E. was also supported by a Stanford Cancer Institute Postdoctoral Fellowship from the Ellie Guardino Research Fund. The saliva sample was obtained from a joint effort between DNA Genotek Inc. and PacBio. We thank Egor Dolzhenko for help visualizing tandem repeats, Adam English for advice on benchmarking TRs, Ed Esplin for the curated list of colorectal adenocarcinoma genes, Raushun Kirtikar and Maha Razzaq for early help with the analysis, and Alexander Honkala for help with text.

## Author contributions

G.S.E. conceived and designed the study. G.S.E. and M.P.S. supervised the research. K.W., H.L., and L.F. led the bioinformatic analyses. C.J.A. led the experimental work, including plasmid construction, cell culture, and luciferase assays. K.Z., W.J.R., and S.B.K. assisted with bioinformatic analyses. P.L. and G.S.E. performed library preparation. J.R.C. constructed plasmids. A.M.H. provided tissue samples. G.S.E., K.W., C.J.A., H.L., and L.F. wrote the manuscript, with input and approval from all authors. G.S.E. and M.P.S. provided funding.

