## Supplemental Information for "Whole-genome variant detection in long-read sequencing data from ultra-low input patient samples"

Supplementary Information

Katherine Wang,[1,2,*] Cera J. Aex,[1,*] Hayan Lee,[2,3,*] Lucas Finot,[1] Kevin Zhu,[2] Julianna R. Chang,[2] Aaron M. Horning,[2] William J. Rowell,[4] Philip Li,[4] Sarah B. Kingan,[4] Michael P. Snyder[2], Graham S. Erwin[1]

[1]Department of Molecular and Human Genetics, Baylor College of Medicine, Houston, TX, USA.
[2]Department of Genetics, Stanford University, Palo Alto, CA, USA.
[3]Present address: Cancer Epigenetics Institute, Nuclear Dynamics and Cancer Program, Fox Chase Cancer Center, Philadelphia, PA, USA.
[4]Pacific Biosciences, Menlo Park, CA, USA.

*These authors contributed equally.

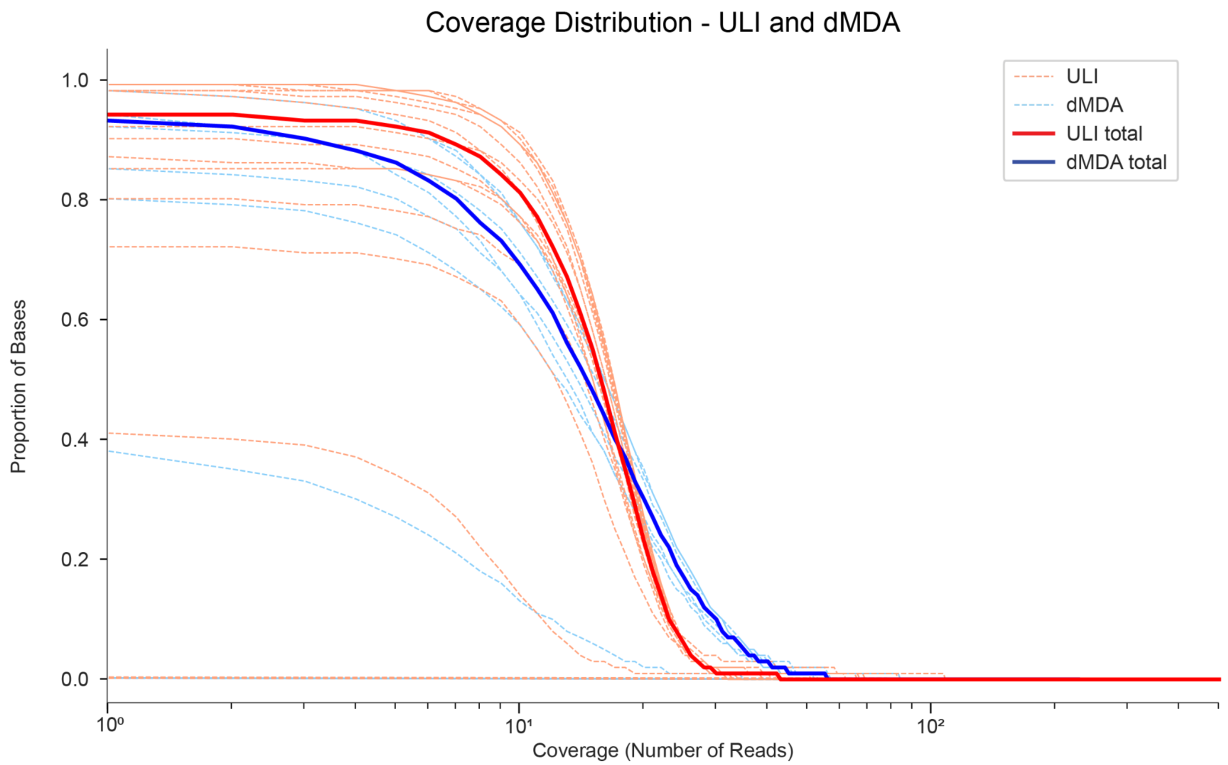

**Figure S1. Standard quality control plot of coverage distribution comparison between NA24385 ULI-HiFi and dMDA sequencing methods.** Both methods shown at an average sequencing depth of 15.5×.

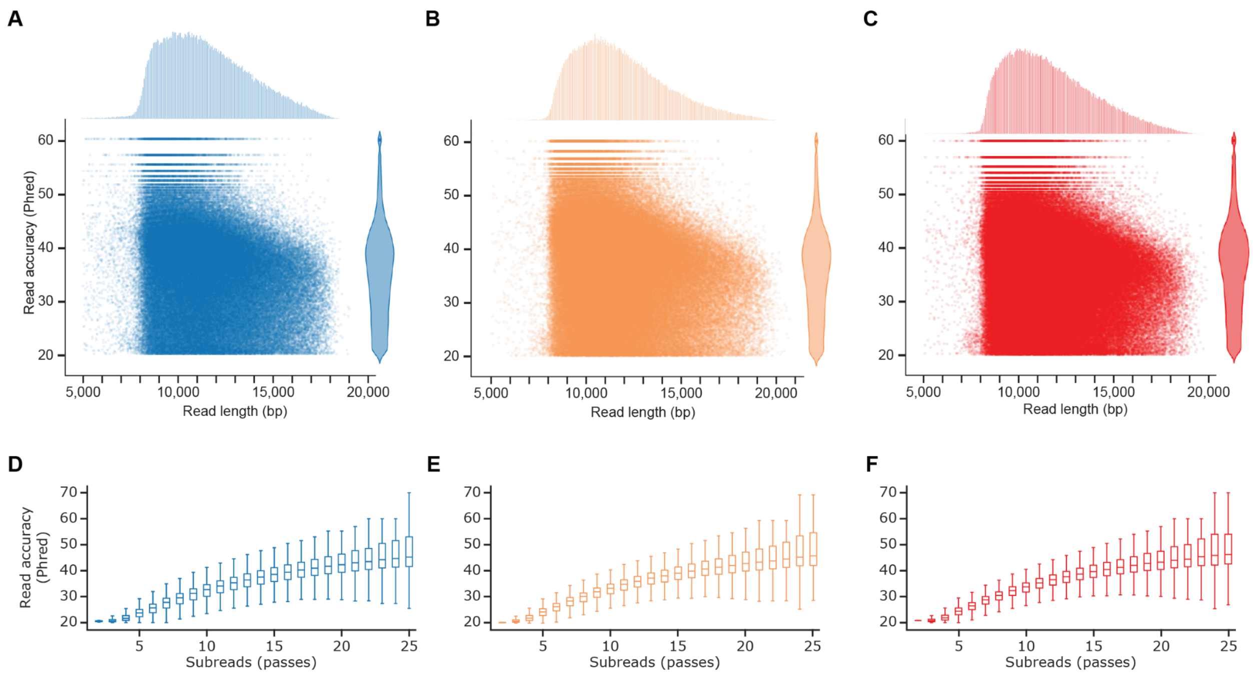

**Figure S2. Accuracy and read length of sequencing data from a patient with familial adenomatous polyposis (FAP). A, B, C)** Sequencing read length (bp) and predicted accuracy (Phred quality score) of adjacent normal (A), polyp (B), and adenocarcinoma (C) reads. **D, E, F)** Accuracy of reads with number of passes, predicted by HiFi CCS software of adjacent normal (A), polyp (B), and adenocarcinoma (C) reads.

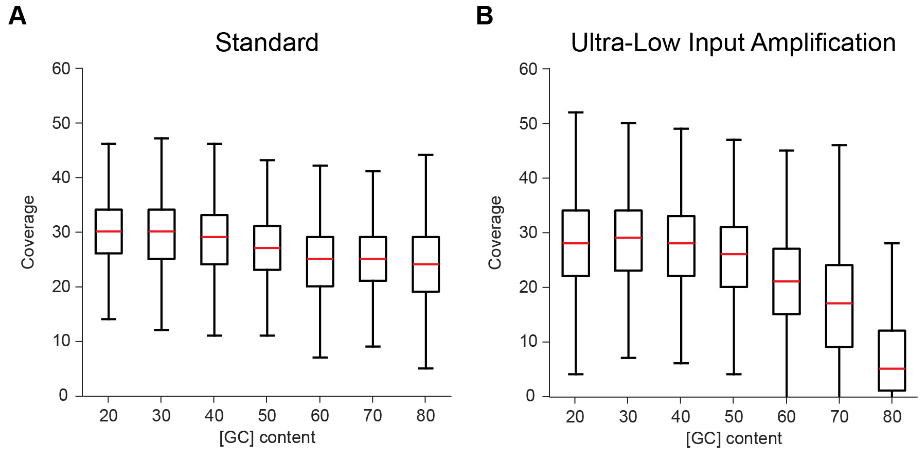

**Figure S3. Coverage across windows of GC content. A)** Standard HiFi coverage across GC content. **B)** ULI-HiFi coverage across GC content**.**

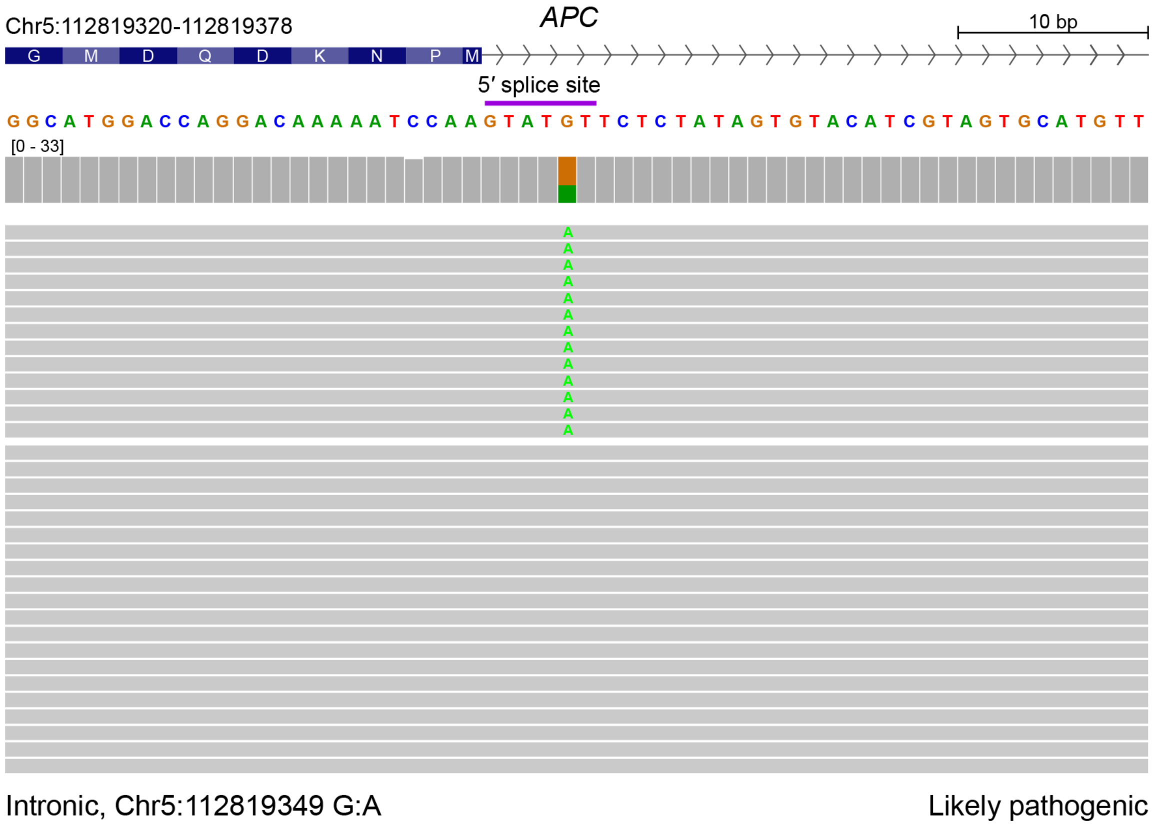

**Figure S4. Detection of the likely pathogenic *APC* mutation in a patient with familial adenomatous polyposis (FAP).** ULI HiFi reads from adjacent normal colon tissue showing a likely pathogenic germline mutation at a 5' splice site in APC.

**Supplemental Table S1.** Benchmarking variant-calling performance across PacBio Revio and Sequel II platforms. Performance of small variant calling (SNVs and indels) was from GRCh38 and measured against Genome in a Bottle (GIAB) small variant reference set v4.2.1. Performance of SV calling was from hs37d5 and measured against GIAB SV reference set v0.6. Performance of TR calling was from GRCh38 and benchmarked with GIAB tandem repeat (TR) reference set v1.0.1. F1 score was calculated as $\frac{2 \times precision \times recall}{precision + recall}$, with an F1 score of 100% indicating perfect precision and recall. All samples were analyzed with DeepVariant v1.4 (SNVs and indels), Sniffles v2.2 (SVs), and TRGT v1.1.1 (TRs).

| | | | **SNVs** | | | **INDELs** | | | **SVs (hs37d5)** | | | **TRs** | | |
|---|---|---|---|---|---|---|---|---|---|---|---|---|---|---|
| **Sample** | **Platform** | **GRCh38 Coverage** | **Precision (%)** | **Recall (%)** | **F1 (%)** | **Precision (%)** | **Recall (%)** | **F1 (%)** | **Precision (%)** | **Recall (%)** | **F1 (%)** | **Precision (%)** | **Recall (%)** | **F1 (%)** |
| NA24385 Revio | PacBio Revio | 15.5× | 99.87 | 99.48 | 99.67 | 94.57 | 95.18 | 94.87 | 93.58 | 93.63 | 93.61 | 95.54 | 97.06 | 96.3 |
| NA24385 Sequel II | PacBio Sequel II | 15.5× | 99.92 | 99.64 | 99.78 | 97.80 | 97.16 | 97.48 | 93.40 | 90.71 | 92.03 | 95.64 | 97.54 | 96.58 |

**Supplemental Table S2.** Quality control statistics, variant counts, and SMRT cell sequencing metrics for NA24385 across three library preparation methods: dMDA, ULI, and Standard HiFi.

| | dMDA | | | | ULI | Standard HiFi 28× | | |
|---|---|---|---|---|---|---|---|---|
| **Indels Called** | 745,701 | | | | 1,929,734 | 1,991,660 | | |
| **SNVs Called** | 3,590,484 | | | | 7,302,657 | 7,126,166 | | |
| **SVs Called** | 81,041 | | | | 32,825 | 31,965 | | |
| **Mean Coverage Depth** | 15.5× | | | | 27.5× | 28.0× | | |
| **Sequencing Platform** | PacBio Sequel II | | | | PacBio Revio | PacBio Sequel II | | |
| | **dMDA Cell 1** | **dMDA Cell 2** | **dMDA Cell 3** | **dMDA Cell 4** | **ULI Cell** | **Cell 1** | **Cell 2** | **Cell 3** |
| **Mean SMRT Cell Read Length (bp)** | 14,549 | 11,726 | 12,604 | 13,456 | 9,385 | 12,852 | 12,856 | 12,862 |
| **Mean SMRT Cell Read Quality (Phred)** | 31.7 | 31.9 | 32.4 | 31.9 | 40.8 | 32.6 | 32.8 | 33.7 |

**Supplemental Table S3.** NA24385 indel benchmarking results with homopolymers removed. Homopolymers, defined as regions with at least 3 consecutive base pairs, were excluded due to their known susceptibility to sequencing and alignment errors in long-read technologies. Performance was from GRCh38 and measured against GIAB small variant reference set v4.2.1.

| | | | | Indels (homopolymers removed) | | |
|---|---|---|---|---|---|---|
| **Platform** | **Sample** | **GRCh38 Coverage** | **Variant caller** | **Precision (%)** | **Recall (%)** | **F1 (%)** |
| PacBio Sequel II | NA24385 Standard | 28× | DeepVariant 1.4 | 99.25 | 99.46 | 99.35 |
| PacBio Revio | NA24385 ULI | 27.5× | DeepVariant 1.4 | 95.32 | 96.74 | 96.02 |
| PacBio Sequel II | NA24385 dMDA | 15.5× | DeepVariant 1.4 | 86.38 | 73.76 | 79.57 |

**Supplemental Table S4. Reproducibility of variant-calling accuracy with downsampling.** Random downsampling to 15.5× performed across three independent replicates for Standard HiFi and ULI-HiFi NA24385. Performance of small variant calling (SNVs and indels) was from GRCh38 and measured against Genome in a Bottle (GIAB) small variant reference set v4.2.1. Performance of SV calling was from hs37d5 and measured against GIAB SV reference set v0.6. Performance of TR calling was from GRCh38 and benchmarked with GIAB tandem repeat (TR) reference set v1.0.1. F1 score was calculated as $\frac{2 \times precision \times recall}{precision + recall}$, with an F1 score of 100% indicating perfect precision and recall. All samples were analyzed with DeepVariant v1.4 (SNVs and indels), Sniffles v2.2 (SVs), and TRGT v1.1.1 (TRs).

| | | | **SNVs** | | | **Indels** | | | **SVs (hs37d5)** | | | **TRs** | | |
|---|---|---|---|---|---|---|---|---|---|---|---|---|---|---|
| **Sample** | **Platform** | **GRCh38 Coverage** | **Precision(%)** | **Recall (%)** | **F1 (%)** | **Precision (%)** | **Recall (%)** | **F1 (%)** | **Precision (%)** | **Recall (%)** | **F1 (%)** | **Precision (%)** | **Recall (%)** | **F1 (%)** |
| NA24385 Standard Sample 1 | PacBio Sequel II | 15.5× | 99.91 | 99.52 | 99.72 | 97.37 | 96.60 | 96.98 | 90.33 | 92.18 | 91.25 | 95.71 | 97.23 | 96.46 |
| NA24385 Standard Sample 2 | PacBio Sequel II | 15.5× | 99.91 | 99.54 | 99.72 | 97.41 | 96.61 | 97.01 | 90.12 | 92.17 | 91.13 | 95.64 | 97.23 | 96.43 |
| NA24385 Standard Sample 3 | PacBio Sequel II | 15.5× | 99.90 | 99.53 | 99.71 | 97.39 | 96.57 | 96.98 | 90.27 | 92.23 | 91.24 | 95.58 | 97.16 | 96.36 |
| NA24385 ULI Sample 1 | PacBio Revio | 15.5× | 99.69 | 98.35 | 99.02 | 92.38 | 91.10 | 91.74 | 87.16 | 86.43 | 86.79 | 90.46 | 91.48 | 90.97 |
| NA24385 ULI Sample 2 | PacBio Revio | 15.5× | 99.69 | 98.35 | 99.01 | 92.34 | 91.13 | 91.73 | 87.25 | 86.81 | 87.03 | 90.51 | 91.5 | 91 |
| NA24385 ULI Sample 3 | PacBio Revio | 15.5× | 99.79 | 98.94 | 99.36 | 90.13 | 91.42 | 90.77 | 87.37 | 85.80 | 86.58 | 87.95 | 93.21 | 90.51 |

**Supplemental Table S5:** Correctable mistakes identified in the NA24385 GIAB/T2T SV reference set from GRCh38. Columns include genomic coordinates (GRCh38), SV type, variant length (bp; deletions notated as negative length, insertions as positive), and gene annotation. Entries labeled "N/A" indicate no gene annotation.

| **Chromosome** | **Start** | **End** | **SV Type** | **SV Length** | **GENE** |
|---|---|---|---|---|---|
| Chr2 | 65,939,407 | 65,939,457 | DEL | -50 | *LINC02934* |
| Chr6 | 167,162,352 | 167,162,432 | DEL | -80 | N/A |
| Chr11 | 1,431,224 | 1,431,307 | DEL | -83 | *BRSK2* |
| Chr4 | 141,283,454 | 141,283,352 | INS | 102 | N/A |
| Chr4 | 190,329,328 | 190,329,276 | INS | 52 | N/A |
| Chr13 | 112,993,783 | 112,993,456 | INS | 327 | *MCF2L* |
| Chr18 | 53,029,668 | 53,029,586 | INS | 82 | *DCC* |
| ChrX | 67,035,047 | 67,034,780 | INS | 267 | N/A |

**Supplemental Table S6.** Coverage of medically-relevant genes using NA24385 ULI-HiFi, dMDA, and SRS methods. Genomic coordinates across publications refer to GRCh38. Average genome-wide coverage of the ULI and dMDA datasets were 27.7× and 15.5×, respectively. Average coverage of the HG002 SRS dataset (GIAB Illumina 2×250) is reported as 40-50×. Lists of medically-relevant genes were previously published in Mandelker et al. 2016 and Wagner et al. 2022.

See attachment.

**Supplemental Table S7. Cancer types with *LIMD1* expansions in the PCAWG dataset.** Tier 1 included clear expansions with strong read coverage, while Tier 2 comprised cases with ambiguous alignments or potential allele dropout. Counts are reported by primary cancer tissue, with totals representing the sum of Tier 1 and Tier 2 events.

| Cancer tissue | Tier 1 Expansion Count | Tier 2 Expansion Count | Total count |
|---|---|---|---|
| Pancreas | 7 | 7 | 14 |
| Central nervous system | 8 | 0 | 8 |
| Lung | 4 | 3 | 7 |
| Kidney | 2 | 4 | 6 |
| Lymph | 5 | 1 | 6 |
| Breast | 3 | 2 | 5 |
| Ovary | 5 | 0 | 5 |
| Liver | 3 | 1 | 4 |
| Prostate | 2 | 2 | 4 |
| Skin | 2 | 2 | 4 |
| Bone | 3 | 0 | 3 |
| Head and neck | 2 | 0 | 2 |
| Stomach | 1 | 1 | 2 |
| Bladder | 1 | 0 | 1 |
| Esophagus | 0 | 1 | 1 |
| Thyroid | 0 | 1 | 1 |
| Uterus | 1 | 0 | 1 |
| **Total** | **49** | **25** | **74** |
